# WebGWAS: A web server for instant GWAS on arbitrary phenotypes

**DOI:** 10.1101/2024.12.11.24318870

**Authors:** Michael Zietz, Undina Gisladottir, Kathleen LaRow Brown, Nicholas P. Tatonetti

## Abstract

Complex disease genetics is a key area of research for reducing disease and improving human health. Genome-wide association studies (GWAS) help in this research by identifying regions of the genome that contribute to complex disease risk. However, GWAS are computationally intensive and require access to individual-level genetic and health information, which presents concerns about privacy and imposes costs on researchers seeking to study complex diseases. Publicly released pan-biobank GWAS summary statistics provide immediate access to results for a subset of phenotypes, but they do not inform about all phenotypes or hand-crafted phenotype definitions, which are often more relevant to study. Here, we present WebGWAS, a new tool that allows researchers to obtain GWAS summary statistics for a phenotype of interest without needing access to individual-level genetic and phenotypic data. Our public web app can be used to study custom phenotype definitions, including inclusion and exclusion criteria, and to produce approximate GWAS summary statistics for that phenotype. WebGWAS computes approximate GWAS summary statistics very quickly (*<*10 seconds), and it does not store private health information. We also show how the statistical approximation underlying WebGWAS can be used to accelerate the computation of multi-phenotype GWAS among correlated phenotypes. Our tool provides a faster approach to GWAS for researchers interested in complex disease, providing approximate summary statistics in short order, without the need to collect, process, and produce GWAS results. Overall, this method advances complex disease research by facilitating more accessible and cost-effective genetic studies using large observational data.

## 1 Introduction

Genome-wide association studies (GWAS) are fundamental to understanding the genetic basis of complex diseases and traits, offering researchers valuable biological insights. However, conducting GWAS requires access to large-scale, individual-level genetic and phenotypic data, such as are provided by the UK Biobank [1] or All of Us [2] projects. Accessing and analyzing such data poses significant challenges, including substantial computational resources, access costs, potential privacy concerns due to sensitive personal information, and the necessity for specialized expertise in data processing and statistical analysis.

To mitigate some of these challenges, publicly released pan-biobank GWAS summary statistics have become invaluable resources for the research community [3, 4, 5, 6]. These studies pre-compute and share GWAS summary statistics for thousands of phenotypes, giving researchers immediate access to genetic association results without handling individual-level data. While this approach alleviates some computational burdens and privacy issues, it has notable limitations—results are restricted to a predefined set of phenotypes and cannot easily be used to study custom phenotype definitions that may be more relevant.

To address these limitations, we present WebGWAS, a free, publicly available web application that allows users to define arbitrary phenotypes and rapidly obtain approximate GWAS summary statistics. WebGWAS is built on a statistical approximation we call indirect GWAS, which uses existing summary statistics to rapidly compute GWAS results for user-defined phenotypes, including specific inclusion and exclusion criteria. Researchers can input custom phe-notype definitions and receive approximate GWAS summary statistics in under 10 seconds. Importantly, WebGWAS neither accesses nor stores any private health information, thereby mitigating privacy concerns associated with the use of sensitive individual-level data.

In addition to facilitating rapid GWAS analyses for custom phenotypes, the statistical approximation underlying WebGWAS can be employed to accelerate the computation of multi-phenotype GWAS when phenotypes are correlated. By exploiting the correlations between phenotypes, our method enhances computational efficiency, reducing the time and resources required for large-scale genetic studies across multiple traits.

Overall, WebGWAS provides a faster and more accessible approach to GWAS, lowering barriers for researchers investigating the genetic underpinnings of complex diseases. By eliminating the need for individual-level data access, access fees, and extensive computational resources, WebGWAS empowers researchers to obtain approximate GWAS results immediately, accelerating the pace of exploratory research into complex traits and diseases.

## 2 Results

### 2.1 Overview of WebGWAS

We built WebGWAS.org, a free, public, efficient web app that can rapidly compute approximate GWAS summary statistics. WebGWAS allows users to define arbitrary phenotypes, compute GWAS results, visualize them, and download them. No protected health information (PHI) is stored on the WebGWAS server—only GWAS summary statistics, phenotypic covariance matrices, and anonymized phenotypic data with *k* = 10 anonymity. No user data are collected beyond which phenotype definitions are submitted. At present, WebGWAS contains data for ICD-10 codes and quantitative measurements in the White British subset of the UK Biobank. Figure 1 shows an overview of WebGWAS.

**Figure 1:**
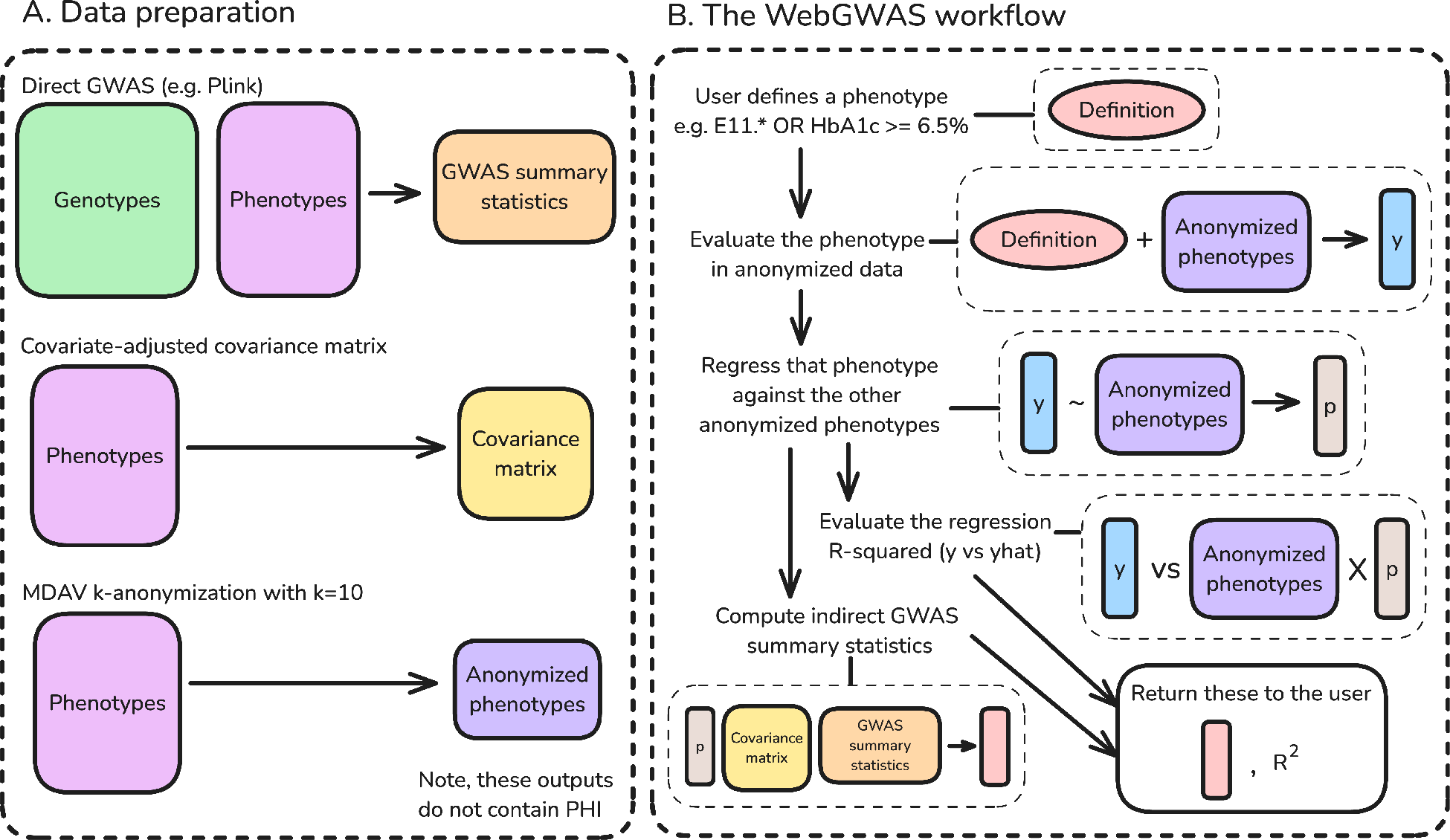
Graphical overview of WebGWAS. **A:** Three artifacts are needed for WebGWAS: GWAS summary statistics, a covariate-adjusted phenotypic covariance matrix, and anonymized phenotypes. No protected health information (PHI) is included in these outputs, which can be uploaded to the WebGWAS server. **B:** Computations on WebGWAS involve five main steps. First, a user defines a phenotype using the web interface. Second, that phenotype definition is evaluated in anonymized data to produce a vector of values. Third, that vector is approximated by the anonymized features using linear regression. Fourth, the fit quality of that linear regression is computed. Fifth, the GWAS summary statistics for the user’s phenotype definition are computed using indirect GWAS. This process only uses the phenotype approximation coefficients, the covariance matrix of the features, and the pre-computed GWAS summary statistics for the features.

### 2.2 Indirect GWAS method

WebGWAS depends on a statistical approach that we term indirect GWAS. The core idea of indirect GWAS is that GWAS summary statistics for a linear combination of phenotypes can be computed without individual-level data, provided certain summary statistics are available for all phenotypes included. For example, a GWAS on the sum of systolic and diastolic blood pressure can be computed given only summary statistics about systolic and diastolic blood pressure, separately. What follows is a high-level overview of this method. A full derivation is provided in the methods.

Consider a genetic association test for a single quantitative phenotype (*y*) and a single genetic variant (*g*), *y ~ g*. Assuming both have zero mean, the coefficient and standard error are the following:

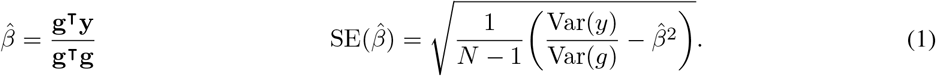

Suppose *y* can be written as a linear combination of *m* feature phenotypes *x*_1_ … *x*_*m*_, (i.e. *y* = ∑*x*_*i*_*p*_*i*_, for some coefficients *p*_1_ … *p*_*m*_). Define two *m*-vectors, **b** and **s**, that hold the coefficient and standard error estimates for regressions of the genetic variant against the feature phenotypes (i.e. *b*_*i*_ = **g**^T^**x**_*i*_*/***g**^T^**g** and *s*_*i*_ = SE(*b*_*i*_)). Let **C** be the phenotypic covariance matrix of the features. Then the coefficient and standard error for *y* can be written in terms of feature summary statistics as

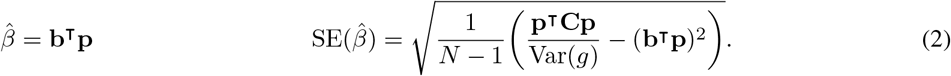

Var(*g*) can be estimated using the coefficients, standard errors, and variances from each feature phenotype and taking the mean across features.

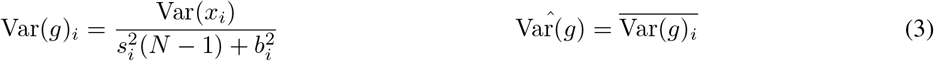

To summarize, the association test summary statistics for phenotype *y* = ∑ *x*_*i*_*p*_*i*_ can be computed using only the coefficients and standard errors for all *x*_*i*_, the covariance matrix among *x*_*i*_, and the coefficients *p*_*i*_. In other words, given appropriate summary statistics about a set of phenotypes, the GWAS summary statistics for any linear combination of those phenotypes can be computed without individual-level data. In the methods we show how this approach is applicable to any linear GWAS method, and how covariates can be included.

### 2.3 Validation of indirect GWAS

To evaluate whether this derivation is correct, we compared to the standard method of direct GWAS on the evaluated linear combination. We used PhenotypeSimulator [7] to simulate genetic and phenotypic data for 10,000 individuals, generated 100 random linear combinations of traits, and compared GWAS summary statistics between direct and indirect approaches to validate our derivation and implementation. The results of this evaluation show that indirect GWAS is mathematically correct in the case of linear models and a good approximation for linear mixed models (Figure 2).

**Figure 2:**
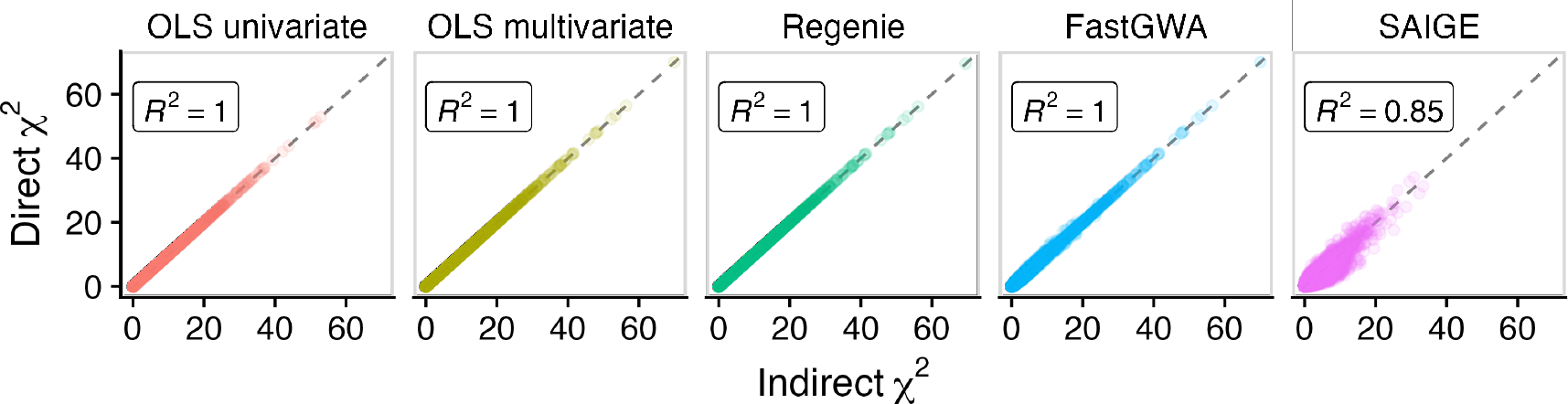
Validation of indirect GWAS using five GWAS methods. GWAS chi-squared statistics are compared between direct and indirect approaches for 100 randomly projected phenotypes, 10,000 genetic variants using simulated data. Each point represents one genetic variant and one phenotype (1,000,000 total points in each facet). *R*^2^ values indicate goodness of fit between the direct and indirect statistics.

### 2.4 Generating summary statistics for arbitrary phenotypes

Research phenotypes are not exclusively defined as linear combinations; they often include boolean operations like “and” and “or”, and inequalities like “greater than”. For example, one could define diabetics as anyone with either an ICD-10 E11 diagnosis or an HbA1c measurement *≥* 6.5%. While arbitrary phenotypes may be nonlinear, indirect GWAS requires them to be linear, so we first linearize each phenotype using linear regression.

Linear approximations perform well for many nonlinear phenotypes, including Phecodes [8] and Boolean operators (Figure 3A). Using ICD-10 code data from the UK Biobank, we generated 140 Phecodes and 300 random nonlinear Boolean combinations, and we approximated these using linear regression against ICD-10 codes. The quality of linear approximations varied greatly between phenotypes. Pairs of codes combined like “x and not y” or “x or y” were the best approximated, with median *R*^2^ values of 0.96 and 0.98, respectively. Phecode performance ranged widely, with regression *R*^2^ values ranging from 0.15 to 0.98, but overall they performed well with a median of 0.82. We found consistently poor performance among phenotypes defined as “x and y”, with *R*^2^ values between 0.01 and 0.39. In short, many nonlinear phenotypes could be approximated with reasonable performance using a linear approximation (74% (324/440) above 0.5, 66% (289/440) above 0.75).

**Figure 3:**
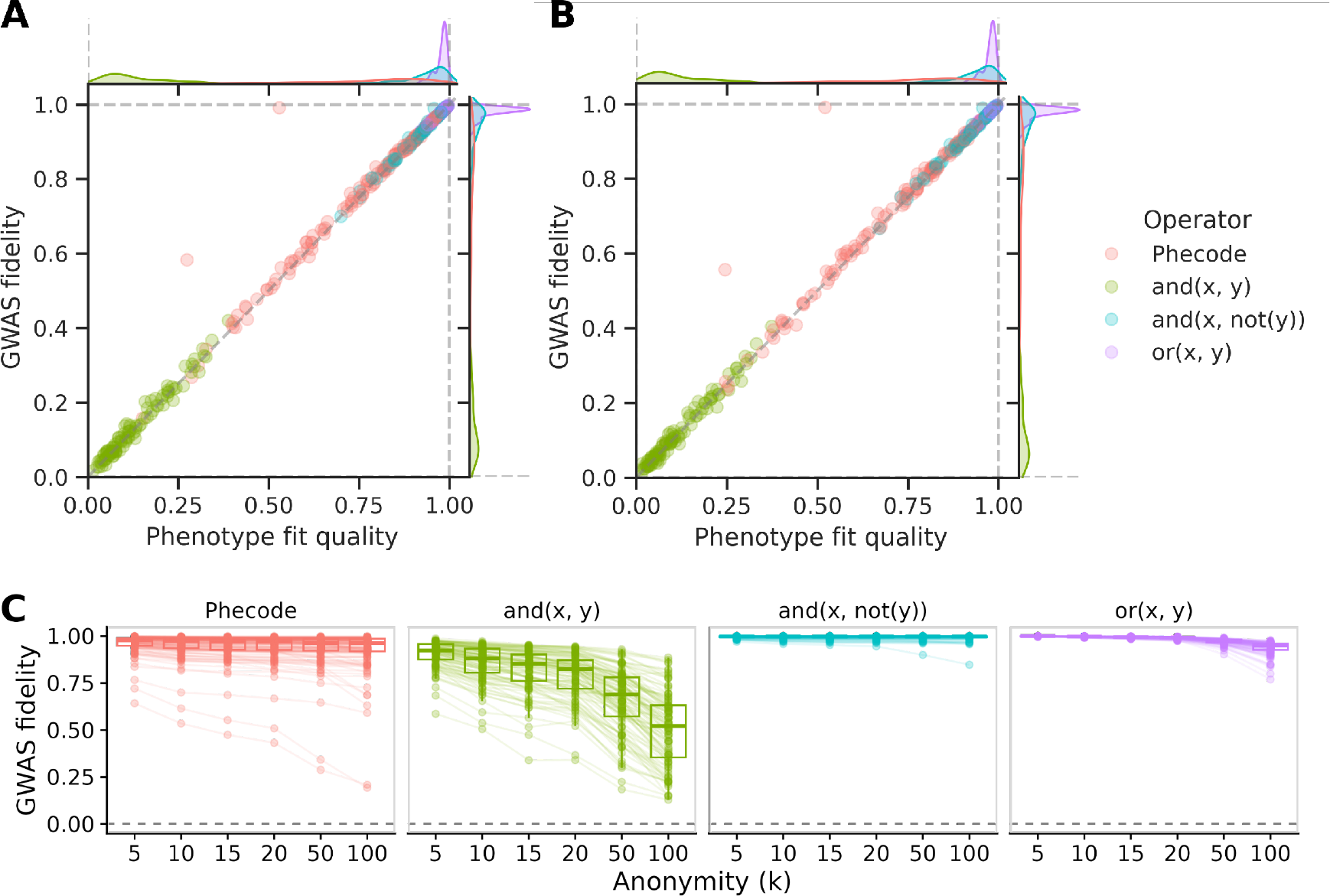
Indirect approach maintains GWAS fidelity for many nonlinear phenotypes. **A:** Phenotype fit quality predicts GWAS fidelity. Nonlinear phenotypes (140 Phecodes and 300 boolean definitions) were approximated using linear combinations of ICD-10 codes via OLS. GWAS fidelity (Pearson *R* between direct and indirect chi-squared statistics) is worsened by linearization alone, and it is highly correlated (Pearson correlation 0.99) with phenotype fit quality (*R*^2^ between exact and linearly approximated phenotypes). Outliers above the line of unity are inflated due to exceptionally high association statistics (see supplementary materials). **B:** Anonymization degrades performance but phenotype fit quality is still highly correlated with GWAS fidelity (Pearson correlation 0.76). ICD-10 code data were first anonymized using MDAV with *k* = 10. Nonlinear phenotypes were evaluated on the anonymized data, then the best linear combinations were estimated in anonymized rather than original data. GWAS fidelity is worsened both due to linearization and anonymization. **C:** Increasing privacy degrades GWAS fidelity. ICD-10 data were anonymized using MDAV at various degrees of anonymity. In each anonymized dataset, all nonlinear phenotype definitions were evaluated, linearized, and used to perform indirect GWAS. GWAS fidelity in this subplot refers to the Pearson *R* between anonymized and non-anonymized indirect GWAS chi-squared statistics. This definition intentionally differs from the previous subplots to isolate the effect of anonymization.

GWAS fidelity is the key metric for evaluating WebGWAS. We quantified GWAS fidelity using the Pearson correlation between direct and indirect GWAS chi-squared statistics. While GWAS fidelity is the metric of greatest interest, it cannot be computed in WebGWAS, because it requires direct GWAS summary statistics. However, we found that phenotypic fit quality (*R*^2^ as above) is an excellent predictor of GWAS fidelity (Pearson correlation 0.99; Figure 3A).

This analysis led us to draw two conclusions. First, many real phenotypes (e.g. Phecodes) can be well-approximated using linear regression (e.g. against ICD-10 codes). Second, phenotypic fit quality—which can be computed in WebGWAS—is an acceptable gauge for GWAS fidelity. In summary, indirect GWAS is applicable to arbitrary phenotypes, and phenotype fit quality, which can be computed cheaply, provides a quality gauge for the results.

### 2.5 Protecting privacy by anonymizing phenotype data

Phenotypic data are used for one purpose in WebGWAS: to find the linear combination of features that best predicts the user-defined phenotype. We hypothesized that anonymized data would give similar coefficients to non-anonymized data, while reducing the risk of exposing sensitive information. To anonymize phenotype data, we chose maximum distance to the average vector (MDAV), an anonymization method that ensures every record is at least *k*-anonymous in the dataset.

Anonymization involves a loss of fidelity. We evaluated whether this loss is empirically tolerable for WebGWAS. We anonymized ICD-10 code data from the UK Biobank to *k* = 10, applied the previous phenotype definitions (i.e. Phecodes and Booleans) to the anonymized data, regressed anonymized nonlinear phenotypes against anonymized ICD-10 codes, then used those coefficients to compute indirect GWAS using GWAS summary statistics from the non-anonymzied ICD-10 codes. Both sources of noise (linearization and anonymization) are present in these GWAS summary statistics. Anonymization worsened the phenotype fit quality (*R*^2^) of linearized phenotypes from 0.71 to 0.37 (Figure 3B). As in the non-anonymized analysis, phenotype fit quality predicted GWAS fidelity (Pearson correlation 0.76), and only phenotypes with fit quality above 0.5 lead to GWAS fidelity above zero. Overall, GWAS fidelity was high for a large fraction of nonlinear phenotypes (65% (285/440) above 0.5, 60% (265/440) above 0.75), indicating that indirect GWAS is applicable even with the added noise from anonymization.

WebGWAS uses *k* = 10 to provide reasonable performance with minimal privacy risk. We evaluated how our other choices of *k* affect GWAS fidelity. This comparison used the same sets of phenotypes (300 Boolean combinations and 140 Phecodes) and features (340 ICD-10 codes) as before. Here, we compared GWAS fidelity of anonymized, linearly approximated data relative to non-anonymized, linearly approximated data in order to isolate the effect of anonymization. We found, unsurprisingly, that GWAS fidelity degraded as the degree of anonymity increased (Figure 3C). With *k* = 10, all phenotypes had a GWAS fidelity above zero, and this decreased with *k* across all phenotypes.

### 2.6 Interactive web app

WebGWAS provides two interfaces for defining phenotypes. The list interface allows users to define phenotypes using ICD-10 codes and two operators, AND and NOT (Figure 4A). The tree interface allows users to define arbitrary phenotypes using ICD codes and quantitative phenotypes, as well as 10 operators (e.g. less than, OR, etc.; Figure 4B). After a phenotype is submitted, a quality gauge appears which shows the phenotype approximation quality, which proxies GWAS fidelity. Finally, once the GWAS summary statistics are computed, an interactive Manhattan plot is shown, and a download link to the full summary statistics appears (Figure 4C).

**Figure 4:**
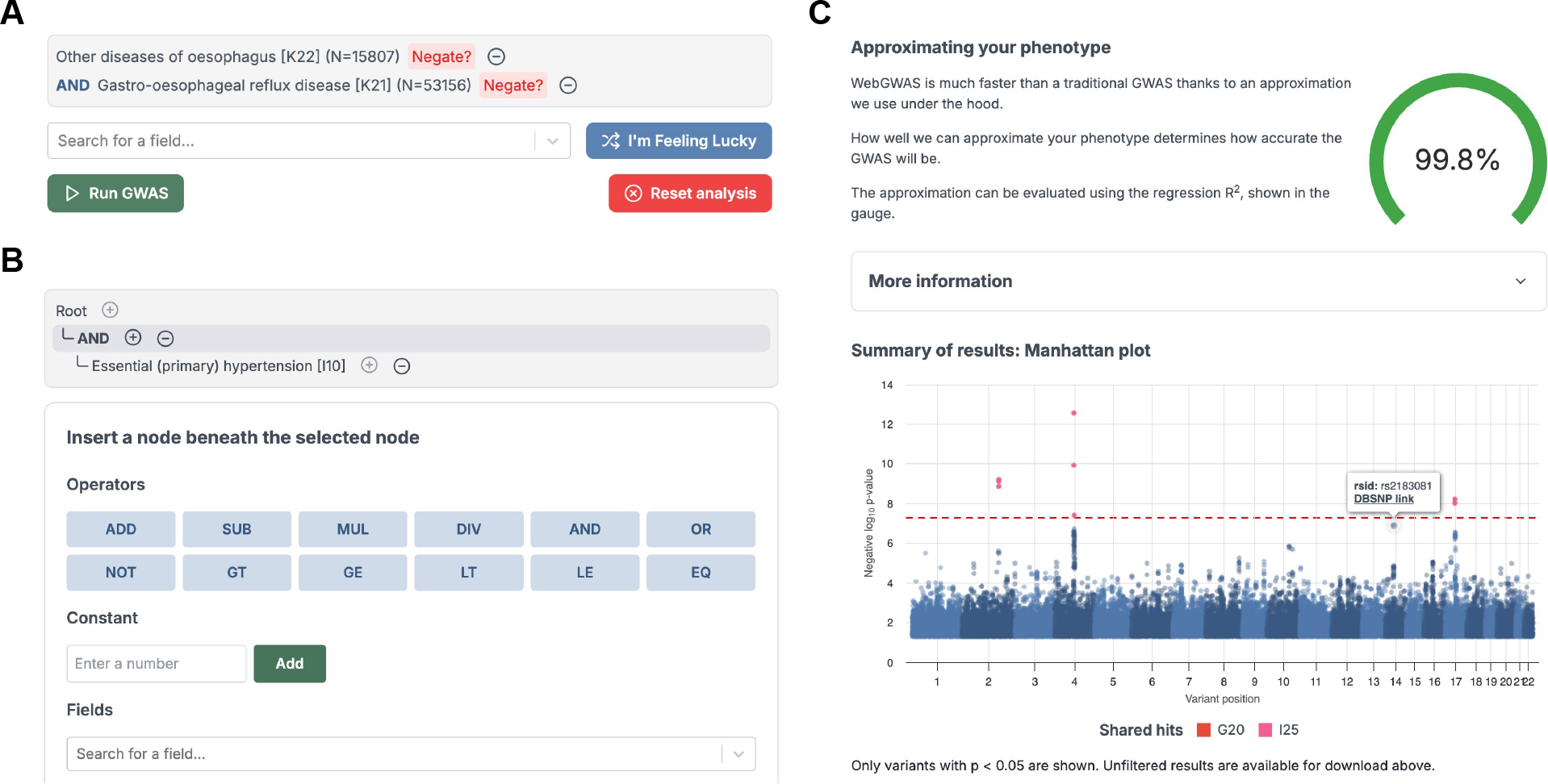
Screenshots of the WebGWAS interface. **A:** The list interface allows users to pick series of ICD-10 codes that define a phenotype using “and” and “not” operations. **B:** The tree interface enables defining truly arbitrary phenotypes using 10 unary and binary operators, constants, and both continuous (e.g. height) and binary (e.g. ICD-10 codes) phenotypes. **C:** Summary information is displayed after indirect GWAS is complete. The phenotype quality gauge indicates the *R*^2^ between the anonymized user-defined phenotype and the anonymized, linearized phenotype. An interactive Manhattan plot shows the results of the indirect GWAS, including clickable links to DBSNP for each variant and color highlighting to indicate which variants also reached genome-wide significance for the feature phenotypes included in the user’s definition.

### 2.7 Accelerating pan-biobank GWAS

In addition to WebGWAS, the underlying indirect GWAS method can be used to accelerate pan-biobank GWAS in exchange for a moderate reduction in result fidelity. To do so, we reduce the phenotype dimensionality with principal component analysis (PCA), perform direct GWAS on some fraction of the principal components (PCs), then reconstruct full-dimensional summary statistics using indirect GWAS. By reducing the number of direct GWAS that must be computed, this approach can save a large fraction of computation time.

We found that a large fraction of ICD-10 codes from the UK Biobank could be reconstructed using only a fraction of the PCs, and that performance increased as more PCs were used (Figure 5). Running this analysis in an even larger dataset, we found that GWAS sensitivity and specificity are largely preserved, even with a small fraction of the PCs (Table 1). In other words, while using fewer PCs diminishes overall GWAS fidelity (Figure 5), fidelity is largely maintained for the most extreme associations (Table 1), which are generally of greatest interest.

**Table 1:** Indirect GWAS performance across various dimensionality reductions. This analysis considered 500,000 variants, 342,350 samples, 1238 ICD-10 phenotypes, and used Plink 2 for GWAS. Variants were considered genome-wide significant if their p-values were less than 5×10^−8^. “% of time” indicates what fraction of the full, direct runtime would be needed for the indirect approach. FP, TP, FN, and TN indicate false positive, true positive, false negative, and true negative.

| % of PCs | % of time | FP | TP | FN | TN | Sensitivity | Specificity | Precision | F1 |
| --- | --- | --- | --- | --- | --- | --- | --- | --- | --- |
| 10 | 10.1 | 2015 | 1222 | 24 | 496739 | 0.981 | 0.996 | 0.378 | 0.545 |
| 25 | 24.0 | 1148 | 1756 | 24 | 497072 | 0.987 | 0.998 | 0.605 | 0.750 |
| 50 | 48.2 | 533 | 2234 | 24 | 497209 | 0.989 | 0.999 | 0.807 | 0.889 |
| 75 | 73.5 | 244 | 2354 | 24 | 497378 | 0.990 | 1.000 | 0.906 | 0.946 |
| 90 | 88.6 | 121 | 2553 | 24 | 497302 | 0.991 | 1.000 | 0.955 | 0.972 |
| 100 | 100 | 1 | 2899 | 24 | 497076 | 0.992 | 1.000 | 1.000 | 0.996 |

**Figure 5:**
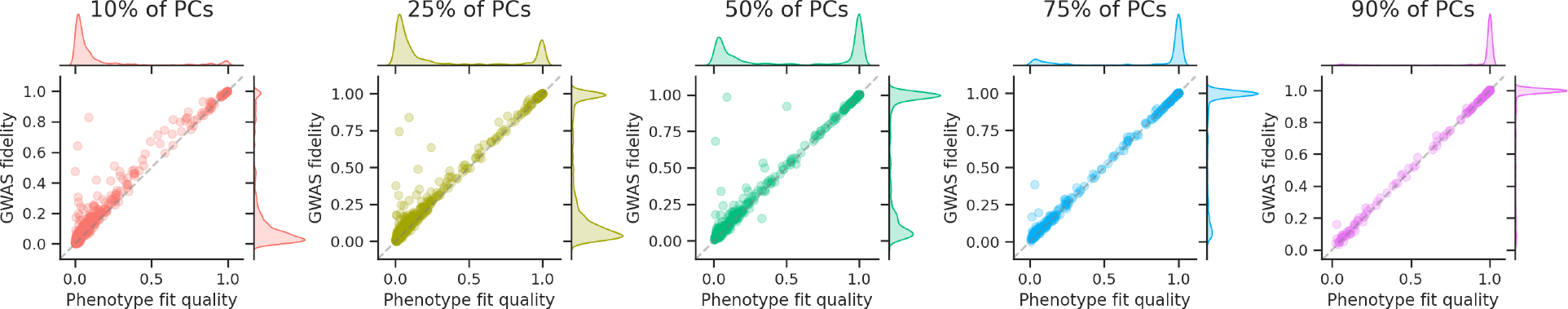
Indirect GWAS with PCA accelerates pan-biobank GWAS. ICD-10 phenotypes from the UK Biobank were compressed to various dimensionalities using PCA. GWAS on the reduced dimensions were used to reconstruct GWAS summary statistics for the original ICD-10 codes using indirect GWAS. Using more PCs improves both phenotype fit quality and GWAS fidelity, and it improves correspondence between the two.

We found that indirect GWAS can substantially reduce computation time for pan-biobank GWAS (Table 1). The time costs for PCA and indirect GWAS are small compared to the runtime savings from computing fewer direct GWAS. In fact, the computation time savings are roughly in line with the latent space fraction used (Table 1), meaning that roughly 50% of computation time can be saved by excluding 50% of PCs. Overall, these results show that indirect GWAS can accelerate pan-biobank GWAS in cases where some degree of approximation is acceptable, and the time savings can be immense.

## 3 Discussion

In this work, we introduced WebGWAS, a free, open source, publicly-available web app for approximate GWAS on arbitrary phenotypes. The underlying method, indirect GWAS, enables computing GWAS for any linear combination of phenotypes, using only GWAS summary statistics and a phenotypic covariance matrix. In order to approximate arbitrary phenotypes in our web app, we paired indirect GWAS with anonymized phenotype data, so that arbitrary phenotypes can be linearly approximated quickly, without exposing PHI. Finally, we showed how indirect GWAS can accelerate pan-biobank GWAS by reducing the number of phenotypes, running GWAS in this reduced dimension, and reconstructing full GWAS results.

WebGWAS has limitations. First, the underlying indirect GWAS method only produces exact results for linear combinations of phenotypes. We showed that linear approximations work well for many real phenotypes, but there remain plenty of phenotypes that are not well approximated. Nonetheless, our approach can quantify the fidelity of its results by measuring the phenotypic goodness-of-fit, thereby providing users with a sense about the accuracy of WebGWAS’s results.

The second major limitation of WebGWAS is that it can only provide GWAS results for linear methods (linear regression and linear mixed models), not logistic regression or generalized linear mixed models. However, linear and logistic regressions produce summary statistics that match closely for variants with small or moderate effects [9], so this limitation is not critical.

Third, WebGWAS can run GWAS on arbitrary phenotypes only insofar as they can be defined in terms of phenotypes with pre-computed summary statistics. For example, a new measurement type that is not already present in the UK Biobank cannot be evaluated with WebGWAS, because it cannot be defined in terms of existing data. Fully out-of-domain phenotypes can be approximated, but doing so is likely to result in poor performance toward the phenotype of interest (see supplementary materials).

Fourth, WebGWAS can only produce results for cohorts, phenotypes, and genetic variants that we have pre-selected and for which we have pre-computed input data (GWAS summary statistics, covariance matrix, and anonymized data). At present, we only include data from the UK Biobank, though we aim to include additional data sets in future updates.

Finally, WebGWAS only produces approximate GWAS summary statistics for nonlinear phenotypes. Even when a user’s phenotype is well-approximated, the GWAS summary statistics are not identical to a direct GWAS using individual-level data. Our web app is intended to be a fast, convenient tool for exploratory analyses. It is not intended to replace a direct approach when producing final research results, which should include thoughtful phenotype definitions, careful quality control, and appropriate GWAS methods. On the other hand, WebGWAS is currently the fastest way to get approximate results without writing code or interacting with data directly and is perfect for exploratory analysis.

We showed how indirect GWAS can be used to accelerate pan-biobank GWAS when approximate results are sufficient. We observed that highly significant associations (low p-values) were generally well-approximated, even at small latent dimensions. As highly significant variants are generally most interesting for further analysis, this suggests that indirect GWAS’s overall lower precision is not critical.

## 4 Methods

### 4.1 Indirect GWAS method

Indirect GWAS is a method to exactly reproduce GWAS summary statistics for a phenotype defined as a linear combination of other phenotypes without directly running a new GWAS. For example, given *m* feature phenotypes *x*_1_,…, *x*_*m*_, define a linear combination *y* =∑ *x* _*i*_*p*_*i*_. Indirect GWAS can reproduce the GWAS results for *y* given only information about the features (*x*_*i*_) and coefficients (*p*_*i*_). To do so, our method needs to compute the following for each genetic variant: coefficient estimate, coefficient standard error, p-value, and sample size. We take the indirect sample size to be the minimum of the sample sizes available for each feature trait, as this is the number of samples that would be available in the equivalent direct approach. We will focus on the coefficient estimate and its standard error, as the p-value can be computed from these statistics.

Consider a regression involving a single genetic variant, a single phenotype, and *N* samples. Let **y** be the *N*-vector of phenotype values, **g** be the *N*-vector of genotype values (coded in any way), and **Z** be the *N × C* matrix of covariates that includes an intercept and is assumed to be full-rank.

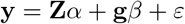

Above, *α* is the *C*-vector of covariate fixed effects, *β* is the scalar effect of the genotype on the phenotype, and *ε* is the *N*-vector of errors. Covariate effects can be removed using a residual projection matrix, **P** = **I**_*N*_ − **Z**(**Z**^T^**Z**)^−1^**Z**^T^. Removing covariates results in residualized phenotype 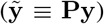, genotype 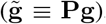, and error 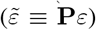 vectors, and it allows writing the regression more compactly.

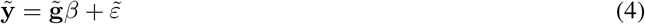

Equation 4 is a univariate least squares regression. Let *d* be the appropriate degrees of freedom for this analysis. The coefficient estimate for this regression is 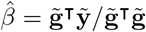, and the standard error is

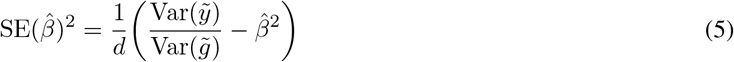

Suppose *y* can be written as a linear combination of *m* feature phenotypes. Let **X** be an *N × m* matrix of feature phenotypes and let **p** be an *m*-vector of the projection coefficients.

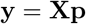

Suppose that for every phenotype in **X**, the same genotype-phenotype regression has been performed. Let **b** and **s** be *m*-vectors holding coefficient estimates and standard errors, respectively, from these regressions (i.e. 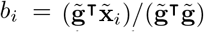 and 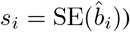). To estimate the standard error for 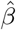, we first need an estimate of 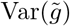, which we obtain by rearranging equation 5 using **b** and **s**.

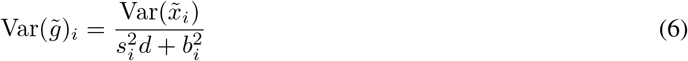

This equation for the genotypic partial variance is written in terms of variables that can be obtained from a GWAS study—the phenotypic partial variance, the estimated coefficient, t he e stimated c oefficient st andard er ror, an d the number of degrees-of-freedom (*d* = *N* − *C* − 1). Every input phenotype *i* may result in a slightly different estimate for this value, so we use the mean across phenotypes as the final estimate for genotype partial variance 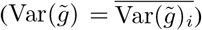. The final term to be estimated is 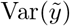, which can be computed using the partial covariance matrix of the feature traits, **C**.

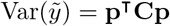

Finally, we can write the coefficient and standard error estimates for *y* as follows:

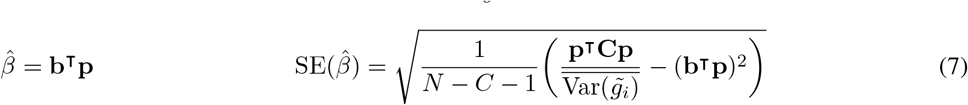

### 4.2 Linear mixed models

Linear mixed models GWAS methods can also be evaluated indirectly. However, the specifics of these operations vary depending on the specifics of the method to be used. In general, linear mixed models are only treated differently when computing the phenotypic partial covariance matrix and when estimating the genotype partial variance. Mixed effect terms should be treated as additional covariates that can be residualized out before computing the phenotypic covariance matrix. This process is simplified for methods like Regenie, which produce intermediate results that include phenotypes residualized against the mixed effect terms. However, some methods (like Regenie) treat chromosomes differently (i.e. they use leave-one-chromosome-out computations), meaning that indirect GWAS needs a different phenotypic covariance matrix for each chromosome. There is no general approach to including linear mixed models, as they all use different approximations and report different intermediate results.

### 4.3 Validation

To confirm that indirect GWAS is mathematically correct, we compared direct vs indirect GWAS summary statistics using simulated data. First, we used PhenotypeSimulator [7] to simulate the following for 10,000 individuals: genotypes for 10,000 genetic variants, 10 phenotypes (*x*_1_ … *x*_1_0), and 5 covariates. Next, we simulated 10 random linear combinations of the phenotypes (e.g. *y*_1_ = 0.5 *× x*_1_ + … + 1.1 *× x*_10_). We performed association tests for all 10 phenotypes against all genetic variants using several different methods: univariate ordinary least squares (OLS; only genotype and intercept), multivariate OLS (genotype, intercept, age, sex, 10 genetic PCs), FastGWA [10] (with covariates), SAIGE [3] (with covariates), and Regenie [11] (with covariates). Finally, we performed association tests on the random projections using both direct and indirect GWAS approaches and compared GWAS chi-squared statistics to verify that indirect GWAS produces equivalent results.

### 4.4 Implementation

Indirect GWAS is mathematically straightforward but challenging to implement efficiently. The method requires potentially thousands of input files, including GWAS summary statistics for every input phenotype, a phenotypic covariance matrix, and a phenotype projection matrix. As the total size of these files can be very large, we provide a high-performance implementation that uses multithreading and chunked processing to provide results efficiently. Our implementation is easily installable, written in Rust, and available on GitHub [12]. We also developed a fast implementation of the data anonymization method (MDAV) [13]. The complete code powering the web application is available on GitHub [14].

### 4.5 UK Biobank data processing

WebGWAS currently contains only data from White British individuals in the UK Biobank. We are working to expand this as soon as possible.

To create the input data to WebGWAS, we first removed individuals whose data were flagged for various reasons as being potentially flawed or erroneous, specifically individuals whose genetic sex was mismatched with their selfreported sex, individuals who were outliers for heterozygosity or missingness (defined by the UK Biobank using the outlier detection algorithm, *abberant* [15]), individuals with ten or more third-degree relatives in the UK Biobank, individuals with sex chromosome aneuploidy, and we restricted to individuals used in the computation of genetic principal components by the UK Biobank. Finally, we restricted to individuals with both binary and quantitative phenotype data available, described below.

For binary phenotypes, we considered all International Statistical Classification of Diseases and Related Health Problems, 10th revision (ICD-10) codes. We mapped and included ICD-10 codes from all six sources available in the UK Biobank: hospital inpatient ICD-9 codes, hospital inpatient ICD-10 codes, self-reported non-cancer illness codes, primary cause of death, secondary cause of death, and general practitioner outpatient diagnoses. Where necessary, we used mappings provided by the UK Biobank to convert each coding to ICD-10. Only codes with at least 100 observations were retained.

For quantitative phenotypes, we considered all quantitative data fields and excluded fields that were not relevant to this study. Specifically, we chose fields meeting the following criteria: only field data (exclude all bulk data, e.g. imaging), unisex fields only, only fields with continuous or integer values, only fields with at least 75,000 samples (per the UK Biobank data dictionary), no strictly genetic fields (e.g. exclude genetic principal components (PCs), exclude pre-computed polygenic scores), no fields dealing with sample quality control (QC) or calibration (e.g. genotyping batch, Affymetrix QC metrics), no fields dealing with home location (e.g. amount of traffic around home location), and no fields dealing strictly with data collection (e.g. the number of samples taken). We manually filtered fields using these criteria.

Many fields have both instance and array indices. Instances correspond to visits to the UK Biobank assessment center. Array indices indicate that multiple measurements or values result from a single investigation instance. We kept array indices as separate fields and took the per-individual mean across instances for each field. Finally, we transformed all quantitative phenotypes using the inverse rank normal transformation, leading to approximately standard normal distributions. Applying the above QC and filtering procedure resulted in 1378 quantitative phenotypes, 1238 binary phenotypes, and 342,350 samples. To save computation time, we restricted our analysis to HapMap3 SNPs, resulting in 1,166,145 SNPs in our final dataset.

## Data Availability

All data are protected health information accessibly through the UK Biobank and cannot be released. Complete processing scripts are available on this project's GitHub.

## 5 Acknowledgments

This work was supported by the NIH NIGMS grant R35GM131905 to Dr. Tatonetti. Access to the UK Biobank was via Approved Research ID 41039. We are grateful to the participants for their generosity in providing their data for research purposes.

## 6 Supplementary methods

### 6.1 Translation between datasets

**Figure S1:**
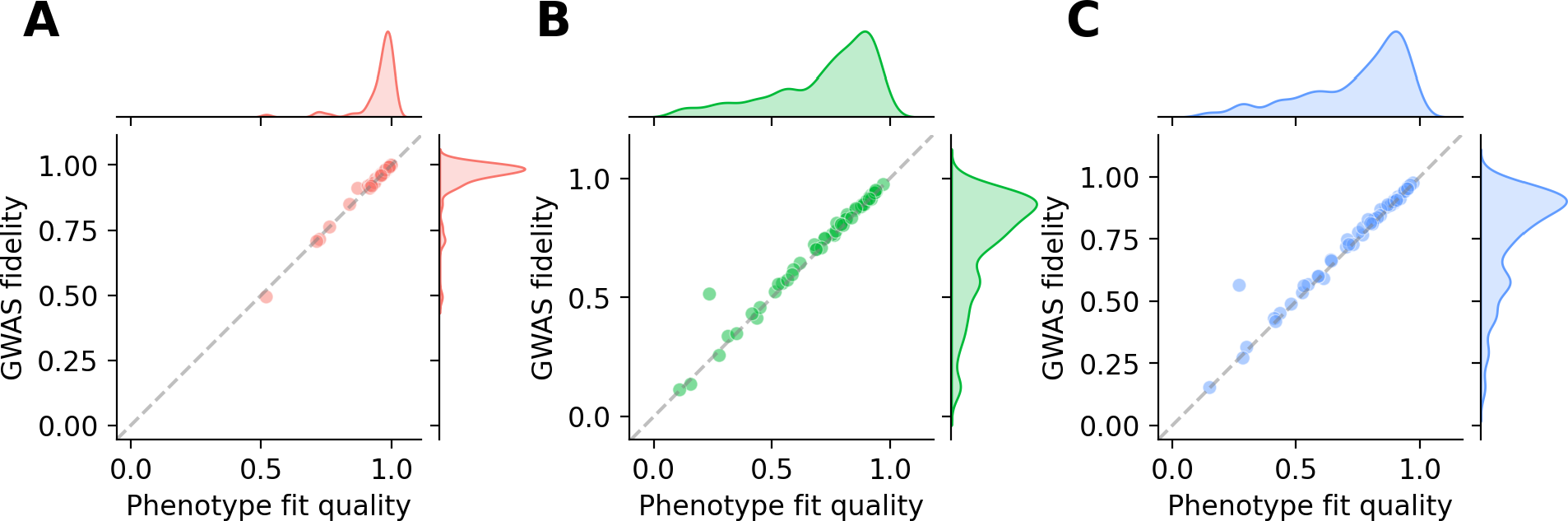
Indirect GWAS can translate between datasets. Phecodes were approximated using ICD-10 codes with linear regression for various training and evaluation datasets. Phenotype approximation coefficients were computed in the training dataset. GWAS fidelity and phenotype fit quality were computed in the evaluation dataset. **A:** Phecode approximations from Cedars-Sinai are similar to UK Biobank approximations. Trained in Cedars-Sinai, evaluated against linear approximations from UK Biobank. **B:** Many Phecode approximations from Cedars-Sinai perform well in UK Biobank as well. Trained in Cedars-Sinai, evaluated against true Phecodes in UK Biobank. **C:** For comparison, approximations from the UK Biobank perform similarly. Linear approximation trained in UK Biobank, evaluated against true Phecodes in UK Biobank.

A limitation of indirect GWAS is that it can only operate on phenotypes defined in terms of its input features (i.e. the phenotypes for which a covariance matrix and GWAS summary statistics are available). In examples presented previously in this paper, the features are ICD-10 codes, meaning that indirect GWAS can only approximate phenotypes that are defined in terms of ICD-10 codes. This is limiting, because there are many phenotypes which might be of interest in GWAS but which do not deal with diagnoses and cannot be explicitly defined as such by a researcher. For example, a researcher interested in genetic associations of UK Biobank questionnaire responses could not explicitly define the response in terms of ICD-10 codes.

However, a large set of real human phenotypes, such as measurements and diagnoses, will span a reasonable fraction of the phenotypes of interest for GWAS. This means that a large dataset like this should have a reasonable amount of statistical predictive power for many real human phenotypes. In some cases, that predictive power could be large enough that a user could reasonably approximate their phenotype of interest using features included in WebGWAS, and use that approximation to gather immediate, approximate GWAS summary statistics. In short, for example, a researcher interested in a particular phenotype could train a statistical or machine learning model to approximate the phenotype using diagnoses and measurements as features, and obtain approximate GWAS summary statistics for that phenotype using WebGWAS. This would be particularly valuable if they could train this model in completely separate phenotype data, for example, an entirely different cohort than the UK Biobank.

Whether this is possible depends on several factors. First, the phenotype of interest needs to be approximable using the features that WebGWAS includes. In building WebGWAS, we attempted to include as many features as possible, to maximize this flexibility for users. However, the space of real human phenotypes is vast, and there are undoubtedly many phenotypes which cannot be approximated well using the features WebGWAS includes. Second, the statistical relationships between the phenotype of interest and the features need to be similar between datasets. For example, even if a user can approximate questionnaire responses using ICD-10 codes and measurements in EHR data from a hospital, this can only translate to good performance in WebGWAS if the correlations among features and the questionnaire are similar in the UK Biobank cohort as in the EHR cohort. This is a fundamental limitation of a cross-dataset approach, and it cannot easily be evaluated statistically without individual-level data from both cohorts.

**Figure S2:**
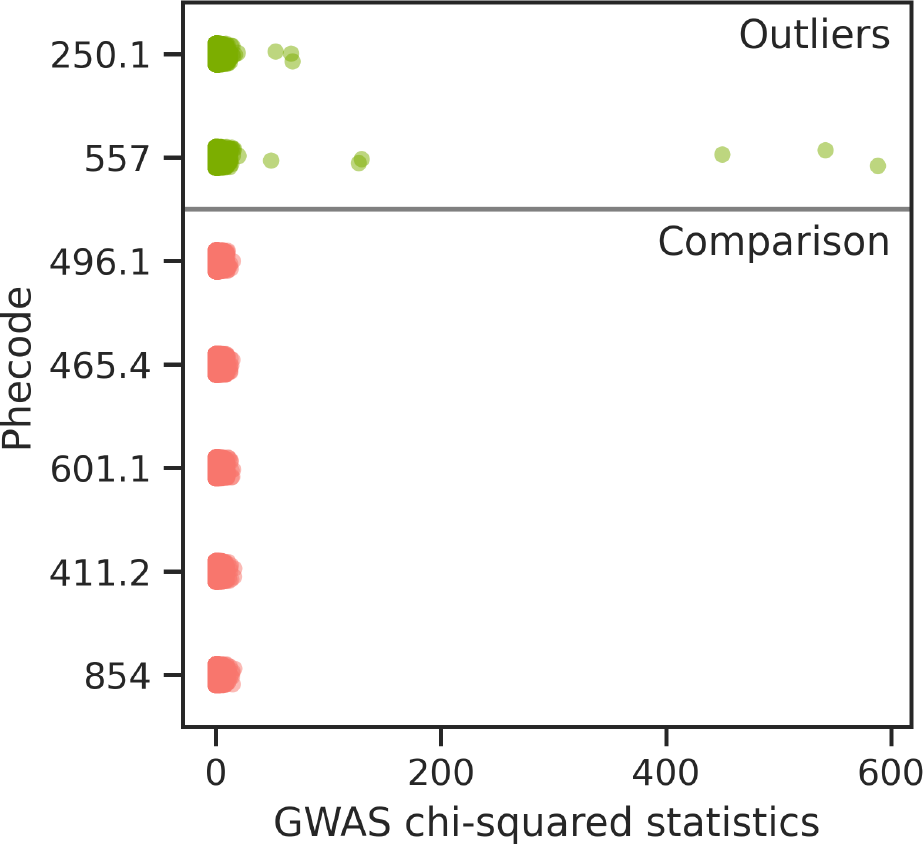
Two outlier phenotypes had moderate phenotype fit quality but exceptional GWAS fidelity, as shown in Figure 3. Upon investigation, the reason for this exceptional GWAS fidelity is that they are outliers in terms of GWAS chi-squared statistics, which leads to inflated variance of chi-squared statistics and inflated Pearson correlation.

In a simple evaluation, we found that Phecode approximations translate very well between a large EHR cohort and the UK Biobank (Figure S1). As features, we used ICD-10 code diagnosis data from Cedars-Sinai Medical Center. We gathered data for 491,822 patients at Cedars-Sinai, and took all 266 ICD-10 codes that had at least 1000 cases in both the UK Biobank and Cedars-Sinai datasets. We then constructed all non-trivial Phecodes that could be constructed with these ICD-10 codes (N=52). In both datasets, we ran linear regressions to approximate the Phecodes using the ICD-10 codes. Finally, we evaluated both regressions in the UK Biobank to obtain three definitions for each Phecode: one exact, one using a UK Biobank linear approximation, and one using a Cedars-Sinai linear approximation.

### 6.2 Investigation of outlier performance

In Figure 3, we noticed that two Phecodes showed GWAS fidelity far above their phenotype fit quality. These two Phecodes were 250.1 (Type 1 diabetes) and 557 (Intestinal malabsorption (non-celiac)). We investigated and found that both were among the lowest prevalence Phecodes in this analysis. Upon further investigation, we discovered that these two phenotypes were outliers in terms of their GWAS summary statistics (Figure S2). Both had a very small number of extreme associations, far beyond the distribution of effects observed for other phenotypes. These outlier variants were also well-approximated with our indirect approach (Figure 2), and they therefore inflated the GWAS fidelity by massively increasing the variance of the chi-squared statistics. Overall, this is an artifact that appears because we used a limited number of genetic variants for this analysis. Our target outcome variable was GWAS fidelity, not genome-wide statistical significance, so we reduced the number of variants that we considered in order to improve computation times. These outliers are in the direction of better fit, not worse, so we believe that they represent a limitation with our evaluation metric (GWAS fidelity) rather than an issue with the method.

## Notes

### Competing Interest Statement

The authors have declared no competing interest.

### Author Declarations

We used only data from the UK Biobank under Approved Research ID 41039.

## References

[1] Clare Bycroft, Colin Freeman, Desislava Petkova, Gavin Band, Lloyd T Elliott, Kevin Sharp, Allan Motyer, Damjan Vukcevic, Olivier Delaneau, Jared O’Connell, et al. The uk biobank resource with deep phenotyping and genomic data. Nature, 562(7726):203–209, 2018.

[2] All of Us Research Program Investigators. The “all of us” research program. New England Journal of Medicine, 381(7):668–676, 2019.

[3] Wei Zhou, Jonas B Nielsen, Lars G Fritsche, Rounak Dey, Maiken E Gabrielsen, Brooke N Wolford, Jonathon LeFaive, Peter VandeHaar, Sarah A Gagliano, Aliya Gifford, et al. Efficiently controlling for case-control imbalance and sample relatedness in large-scale genetic association studies. Nature Genetics, 50(9):1335–1341, 2018.

[4] Pan UKBB Team at Broad Institute. Pan-UK Biobank. pan.ukbb.broadinstitute.org, Accessed: 2023-08-01.

[5] Masahiro Kanai, Masato Akiyama, Atsushi Takahashi, Nana Matoba, Yukihide Momozawa, Masashi Ikeda, Nakao Iwata, Shiro Ikegawa, Makoto Hirata, Koichi Matsuda, et al. Genetic analysis of quantitative traits in the japanese population links cell types to complex human diseases. Nature Genetics, 50(3):390–400, 2018.

[6] Kazuyoshi Ishigaki, Masato Akiyama, Masahiro Kanai, Atsushi Takahashi, Eiryo Kawakami, Hiroki Sugishita, Saori Sakaue, Nana Matoba, Siew-Kee Low, Yukinori Okada, et al. Large-scale genome-wide association study in a japanese population identifies novel susceptibility loci across different diseases. Nature Genetics, 52(7):669– 679, 2020.

[7] Hannah Verena Meyer and Ewan Birney. Phenotypesimulator: A comprehensive framework for simulating multi-trait, multi-locus genotype to phenotype relationships. Bioinformatics, 34(17):2951–2956, 2018.

[8] Joshua C Denny, Marylyn D Ritchie, Melissa A Basford, Jill M Pulley, Lisa Bastarache, Kristin Brown-Gentry, Deede Wang, Dan R Masys, Dan M Roden, and Dana C Crawford. Phewas: demonstrating the feasibility of a phenome-wide scan to discover gene–disease associations. Bioinformatics, 26(9):1205–1210, 2010.

[9] Doug Speed and David J Balding. Sumher better estimates the snp heritability of complex traits from summary statistics. Nature Genetics, 51(2):277–284, 2019.

[10] Longda Jiang, Zhili Zheng, Ting Qi, Kathryn E Kemper, Naomi R Wray, Peter M Visscher, and Jian Yang. A resource-efficient tool for mixed model association analysis of large-scale data. Nature Genetics, 51(12):1749– 1755, 2019.

[11] Joelle Mbatchou, Leland Barnard, Joshua Backman, Anthony Marcketta, Jack A Kosmicki, Andrey Ziyatdinov, Christian Benner, Colm O’Dushlaine, Mathew Barber, Boris Boutkov, et al. Computationally efficient whole-genome regression for quantitative and binary traits. Nature Genetics, 53(7):1097–1103, 2021.

[12] Michael Zietz and Nicholas Tatonetti. Indirect GWAS v0.0.2. 2024. https://github.com/tatonetti-lab/indirect-gwas, 2024.

[13] Michael Zietz. MDAV v0.5.4. 2024. https://github.com/zietzm/mdav, 2024.

[14] Michael Zietz. WebGWAS Backend v0.8.0. 2024. https://github.com/tatonetti-lab/webgwas-backend, 2024.

[15] Céline Bellenguez, Amy Strange, Colin Freeman, Wellcome Trust Case Control Consortium, Peter Donnelly, and Chris CA Spencer. A robust clustering algorithm for identifying problematic samples in genome-wide association studies. Bioinformatics, 28(1):134–135, 2012.

